# Movement disorders ontology for clinically oriented and clinicians-driven data mining of multi-center cohorts in Parkinson’s disease

**DOI:** 10.1101/2020.11.09.20228577

**Authors:** Deepak K. Gupta, Massimo Marano, Raj Aurora, James Boyd, Satya S. Sahoo

## Abstract

Parkinsonian disorders, including Parkinson’s disease (PD) and atypical parkinsonian disorders (APD), are characterized by shared clinical features of parkinsonism and although there are distinct clinical and pathological diagnostic criterion for PD and APD, patients present in the clinic with overlapping clinical features, which evolve with a great deal of variability and complexity over time. This leads to high level of uncertainty in the prediction of diagnosis and progression for an individual patient on clinical grounds. There have been recent initiatives to make available large-scale datasets from multiple research studies such as Accelerating Medicines Partnership for Parkinson’s Disease (AMP PD). However, these is a clear lack of a common terminological system or ontology that can support query analysis of datasets in AMP PD and map across multiple instruments used in the assessment of PD as well as APD. To address these challenges, we developed the Movement Disorder Ontology (MDO) that used a systematic analysis of movement disorder instruments, extensive review of literature led by a movement disorder specialists, and the AMP PD knowledge portal with an iterative ontology engineering process. The current version of MDO is focused on parkinsonian disorders with 203 concepts modeled in three broad categories of: (1) neurological findings, (2) treatment plans, and (3) instruments used to evaluate various traits of PD. MDO holds potential for use in clinical research especially in the context of large-scale phenotypic data available in public repositories such AMP PD with support for concept-based data analysis and potentially correlating with genotypic data.

## Introduction

Movement disorders is a branch of neurology in medicine, and is comprised of several disabling, common and rare neurological disorders, including Parkinson’s disease (PD), essential tremor, dystonia, Huntington’s disease etc. PD is the most well-known of all movement disorders, with 2-4% of the population older than 60 years having PD, with projected increase in prevalence from 7 million cases to 14-17 million by the 2040 worldwide (1). Related to PD, there is a group of rare but rapidly progressive and even more disabling disorders, collectively termed as atypical parkinsonian disorders (APD), including Lewy body dementia, multiple system atrophy, progressive supranuclear palsy, and corticobasal degeneration. On the other hand, essential tremor is the most prevalent and one of the most common movement disorder and neurological disorder, respectively, affecting approximately 2.2% of the US population. Parkinsonian disorders, including PD and APD, are characterized by shared clinical features of parkinsonism, which is further defined as presence of bradykinesia, with rest tremor, rigidity and/or postural instability (2, 3). Although there are distinct clinical and pathological diagnostic criterion for PD and APD, patients present in the clinic with overlapping clinical features, which evolve with a great deal of variability and complexity over time (4). This leads to high level of uncertainty in the prediction of diagnosis and progression for an individual patient on clinical grounds. These challenges faced by patients, their families and clinician are further exacerbated by the lack of sensitive or specific diagnostics tests and biomarkers (5).

Furthermore, the current Movement Disorders Society (MDS) clinical diagnostic criteria for PD attempt to classify patients in two level diagnostic certainty, that is, “clinically established” and “clinically probable PD” (6). This classification is based on the presence of parkinsonism, absence of nine absolute exclusion criteria, presence of at least 2 out of 4 supportive criteria, and absence of red flags or counterbalance of red flags with supportive criterion. Diagnosis of PD is made in clinic by primary care clinicians, neurologists and movement disorders specialists using the presence of key motor features, such as the presence of asymmetric limb bradykinesia with rigidity and/or rest tremor, when accompanied by one or more of the non-motor features (7). In addition, dopamine transporter (DaT) imaging has been recently used to supplement clinical suspicion of PD (8). However, the complexity and overlap of parkinsonian disorders make it extremely difficult to make definite diagnosis in clinic, especially early on., For example, PD patients do not present with a common set of symptoms and signs, and rather have different phenotypes such as with or without rest tremor. In addition, patients with APD often present with clinical features that are similar to PD. Therefore, there is a clear need for a clinical decision support tool that can integrate, and organize relevant data for informed decision making (9).

The availability of the Accelerating Medicine Partnership in Parkinson Disease (AMP PD) database as a high-quality knowledge resource with multi-modal data from 4298 participants presents a unique opportunity to develop new data-driven predictive algorithms for addressing the critical challenges in PD. AMP PD is a unique resource for cohort-based study of different phenotypes and patterns of clinical progression, and biomarkers of PD. In our preliminary work for this research proposal, we analyzed data from AMP PD to characterize the prevalence and patterns of rest and action tremor in PD. However, such conventional single hypothesis-driven approach of using AMP PD database requires manual processing and significant amount of researcher time, which is inherently limited in its approach and impact. To translate the AMP PD database for broader and meaningful discovery of knowledge and applicability to individual patient in a personalized manner on a scalable basis, we need a comprehensive informatics-based approach with machine learning component. There are significant mapping and integration challenges that need to be addressed for integrated analysis of individual patient and AMP PD dataset. A key critical gap in this respect is the lack of a comprehensive domain ontology for movement disorders, which can enable formal knowledge model for data input, semantic data integration, and support analytical queries for predictive modeling in parkinsonian disorders.

Specifically, to support this and similar data mining studies, a movement disorder ontology can serve a critical role as a reference knowledge model to support ontology-based data access (OBDA), which involves mapping of user query terms to database schema, automatic reconciliation of data heterogeneity, and accurate data retrieval (10). Although an ontology for Parkinson’s Disease (PDON) has been developed (11); however, the focus of PDON is on <XYZ> that significantly limits it use in clinical as well as research data query and retrieval in AMP PD and other similar repositories. In this paper, we introduce the Movement Disorder Ontology (MDO) that has been designed to: (1) Improve access to both research and clinical trial data being made available by the AMP PD project and maintained by the Parkinsonian Study Group (PSG) (12); (2) Enable interoperability and mappings between a variety of instrument/scales that are used in the evaluation of patients in movement disorder clinical centers; and (3) Facilitate multi-institution research studies through the development of interoperable data management tools with adequate statistical power. The MDO is also designed to support a systematic evaluation of the different movement disorder instruments in terms of: (1) coverage of movement disorder traits; (2) unique terms measured by each instrument; and (3) objective functions measured by the instrument.

## Background

The challenges associated with the prediction of diagnosis, progression, and prognosis in PD has led to multiple initiatives to provide new insights regarding the underlying causes for the two core traits of PD, namely neuronal death and accumulation of protein α–synuclein in neurons called Lewy bodies. However, there are currently no drugs approved to modify the disease, which highlights a critical need to develop new techniques for biomarker discovery that can integrate phenotype, genomics, proteomics, and related data into predictive models for individual patients. Therefore, a domain-specific ontology for movement disorders is a key knowledge resource to reconcile data heterogeneity, facilitate data access, and support semantics-based knowledge discovery. In this section, we provide an overview of the data resources and technologies used in the development of MDO.

### AMP PD

The AMP initiative brings together the National Institute for Neurological Disorders and Stroke (NINDS), the Food and Drug Administration (FDA), the Michael J. Fox Foundation, and multiple private sector companies, including Verily and Pfizer, to advance research in various disease domains (13). The AMP PD database harmonizes data from four cohort research studies: (1) the BioFIND study; (2) the Harvard Biomarkers study (HBS); (3) the Parkinson’s Disease Biomarker Program (PDBP), and (4) the Parkinson’s Progression Markers Initiative (PPMI). There is a total of 4,298 participants (2,390 males and 1,908 females; 93/8% white / Caucasian, 2.1% Black / African American; age range 20-89) in AMP PD database, including PD patients, 1,476 controls and 293 other subjects (e.g., Multiple System Atrophy, Progressive Supranuclear Palsy, Essential Tremor, Alzheimer’s disease etc.). The BioFIND is MJFF-funded an observational, multi-center cross-sectional cohort of moderate to advanced PD patients on dopaminergic treatment, and contributed 213 subjects (118 PD, 89 controls and 6 others) to AMP PD. The HBS is a biobank of PD and other neurodegenerative disorders, such as Alzheimer’s disease, and contributed 876 subjects (649 PD, 227 controls) to AMP PD. The PDBP is an NINDS-funded, multi-center, longitudinal observation cohort of PD and related disorders of all stages, and contributed 1,599 subjects (882 PD, 549 controls, 168 others) to AMP PD. The PPMI is an MJFF-funded, multi-center, longitudinal observational cohort of early, drug-naïve PD patients, and contributed 1,610 subjects (880 PD, 611 controls and 119 others) to AMP PD. Therefore, AMP PD is a unique resource for cohort-based study of different phenotypes and patterns of clinical progression, and biomarkers.

### Clinical Rating Scales for Parkinson Disorder

Several clinical rating scales have been developed for assessing severity, progression of motor, quality of life and activities of daily living (ADL) in movement disorders(14). Important examples include MDS-Unified Parkinson Disease Rating Scale (MDS-UPDRS), Montreal Cognitive Assessment (MoCA), REM Sleep Behavior Disorder Screening Questionnaire (RBDSQ), the University of Pennsylvania Smell. Identification Test (UPSIT) and the Parkinson’s Disease Questionnaire (PDQ-39). Several of these have been used as primary or secondary outcome measures in clinical trials and cohort studies for PD. In particular, the MDS-UPDRS is one of the most commonly used clinical rating scale for PD, and covers motor and non-motor features of PD in four parts, namely 1) Non-motor Experiences of Daily Living; II: Motor Experiences of Daily Living; III: Motor Examination; IV: Motor Complications.

### Evaluation Protocols used for Diagnosis, Progression, and Prognosis for Movement Disorders

The diagnostic approach to Parkinson’s disease and in general for all movement disorders is based on phenomenology of movements. For example, for PD, the presence of bradykinesia, rigidity and tremor in combination point to a clinical diagnosis of Parkinson’s disease together with a sustained response to dopaminergic agents (i.e. levodopa). The diagnosis should be confirmed by the exclusion of other potential causes of parkinsonism by the history and a detailed examination aimed at confirming the absence of red flags. The latter includes various clinical aspects that would point to diagnosis other than PD (e.g. a prominent dysautonomia with early falls would indicate a multiple system atrophy, while a gaze palsy would suggest a progressive supranuclear palsy). Radiological studies, such as dopamine transporter (DaT) scan are sometimes utilized to assess for evidence of dopaminergic denervation in support of parkinsonian disorder pathology, including PD. There are otherwise no definite biomarkers to confirm clinical diagnosis of PD.

### Related Work

Terms describing movement disorders and PD in particular have been modeled in a variety of biomedical ontologies, such as SNOMED CT and the National Cancer Institute Thesaurus (NCIT); however, we were able to identify only the Parkinson’s Disease Ontology (PDON) as a focused domain-specific ontology for PD (11). The PDON ontology uses a view-based approach to represent terms related to PD, for example “clinical view”, “model of Parkinson’s Disease”, and “familial neurodegenerative disease” among others. Although PDON aims to model a variety of terms ranging from molecular features to clinical concepts with support for text mining and data retrieval, there are several issues in terms of its coverage, ontology modeling approach, and class hierarchy structure that limits its application in clinical research informatics applications. For example, many of the terms in the class hierarchy represent “part of” (partonomy) relation instead of a “is a” (subsumption) relation (“Alpha-synuclein” is modeled as a subclass of “Aggregation_of_alpha-synuclein” instead of using a “part of” relation. Further, the level of granularity required to support ontology-mediated data access as well as querying of large datasets such as AMP PD is not represented in PDON. For example, bradykinesia of different types, such as finger tapping and foot stomping, are modeled at a coarse level of granularity as “Bradykinesia_symptom” in PDON. In contrast, MDO models all the terms related to “rapid alternating movements” at a fine-level of granularity (described in Method section below) to support ontology-mediated query and retrieval of data form AMP PD and other similar datasets. Therefore, after a systematic evaluation of PDON, we decided to develop MDO as a domain-specific ontology for the broader community of movement disorder with sufficient level of details for PD, atypical PD (APD), and other types of movement disorder.

## Method

The development of the ontology was based on three criteria: (1) literature survey on the phenotypic and genotypic characteristics of parkinsonian disorder; (2) diagnostic criteria used for patient evaluation; (3) metadata as well as data attributes of research cohorts, including longitudinal study methods and cohort inclusion exclusion criteria. The ontology was developed using an iterative collaborative process with the use of clinical workflow used by movement disorder specialists as reference protocol for prediction of diagnosis, progression, and prognosis. In the following sections, we describe the structure of the ontology, the application of the MDO terms to AMP PD cohort studies, and the creation of mappings between MDO and AMP PD data for clinical research analysis.

### MDO Structure

During evaluation of a patient, the movement disorder specialist elicits information regarding the patients’ motor symptoms, non-motor symptoms, and ADL together with social as well as family history, which constitute the patient’s perspective on movement disorder related traits. As part of the clinical evaluation, a set of neurological examinations are performed together with standard physical examination of the patient. The neurological examination consists of 15 components that include cognitive functions, rapid alternating movements, involuntary movements, speech abnormality, and gait. The ontology class structure models the different components of the neurological examination at a fine level of granularity that can be directly mapped to both clinical data collected from patients and the cohort study data in AMP PD. For example, Figure 1 illustrates a conceptual view of the three categories of rapid alternating movements modeled in MDO, namely: (1) Bradykinesia; (2) Hypokinesia; and (3) Dysrhythmia. The terms are arranged in a subsumption hierarchy that are modeled as a class-subclass hierarchy in MDO.

**Figure 1:**
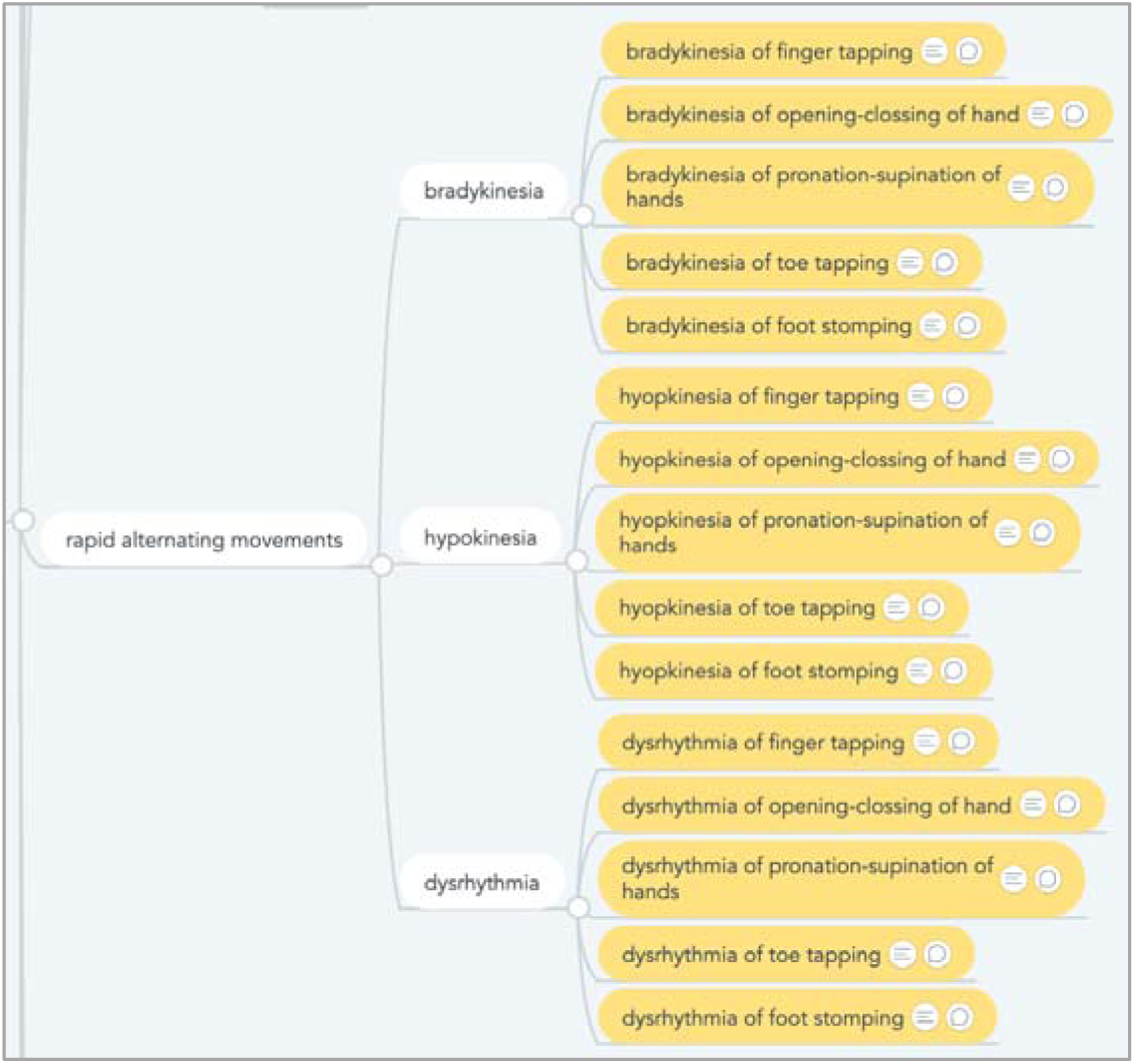
Conceptual view of the terms describing rapid alternating movement

MDO was developed using the Protégé ontology editor and knowledge acquisition system (15) and we used the World Wide Web Consortium (W3C) Web Ontology Language (OWL2), which is based on description logic, to represent MDO (16). The ontology was developed using ontology engineering best practices that included use of structural metrics to evaluate the information content of the ontology and identify modeling errors, and improve the structure of class hierarchy. These structural metrics included maximum depth of the class in the hierarchy, average and maximum number of sibling classes. In addition to the metrics to evaluate the ontology structure, we also used the National Center for Biomedical Ontologies (NCBO) BioPortal to identify and re-use terms from existing ontologies in MDO. For example, we re-used terms from: (1) RxNorm to represent drug ingredients and trade names (17); (2) the Foundational Model of Anatomy (FMA) to model brain anatomy details (18); and (3) the Systematized Nomenclature for Medicine (SNOMED CT) to represent clinical terms (19). The MDO class hierarchy uses the Basic Formal Ontology (BFO) as a reference upper level ontology and uses specific BFO classes to broadly organize the movement disorder terms into the two categories of Occurrent (representing dynamic entities such as a clinical procedure) and Continuant (representing entities that persist through time such as a fluid biomarker) (20). Figure 2 shows the class hierarchy in MDO corresponding to the rapid alternating movements described above. MDO uses multiple OWL annotation properties to represent both human readable description of a class and mappings to existing ontology classes.

**Figure 2:**
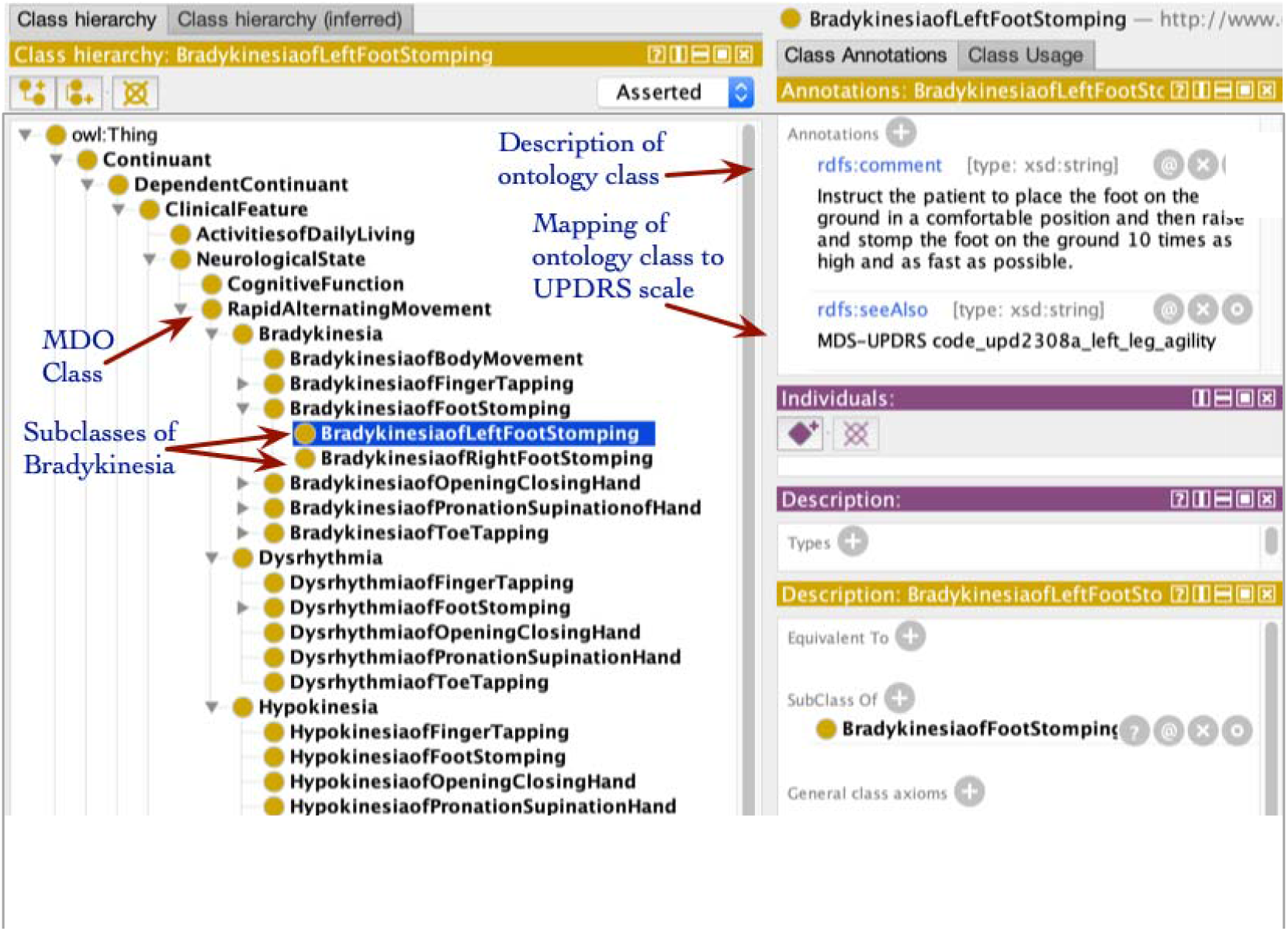
A section of the MDO class hierarchy with classes describing rapid alternating movement with class annotations and mapping to MDS-UPDRS scale

### Mapping to Instruments

One of the primary objectives of MDO is define mappings between the different scales used during patient evaluation and in research cohort studies to facilitate both data access and analysis. We mapped the MDO elements to the MDS-UPDRS scale as an example. Bradykinesia is defined as slowness of movement AND decrement in amplitude or speed (or progressive hesitations/halts) as movements are continued, and is the core feature of PD and its presence is required for clinical diagnosis of PD. Current approach of defining and evaluating for bradykinesia combines three distinct elements into one, specifically, decrease in speed, amplitude bradykinesia (slowness of movements), hypokinesia (decrement in amplitude) and dysrhythmia (progressive hesitations/halts), on different components of the MDS-UPDRS scale, specifically, finger tapping, hand movements (3.5), pronation-supination movements (3.6), toe tapping (3.7), and foot tapping (3.8). However, more often than not, patients have one or more than one of these three elements, and yet, all such patients are classified having bradykinesia. Using MDO, a clinician can record distinct elements of bradykinesia during the clinical examination, which can be then mapped to the MDS-UPDRS scale still without any problem, as well as allow querying of the database approach using bradykinesia with different level of granularities.

## Results

The current version of MDO consists of terms representing the phenotypic traits of movement disorder, the components of diagnostic criteria used during patient evaluation, and data elements used in the AMP PD cohort studies, which are modeled as subclasses of the three BFO classes of *DependentContinuant, IndependentContinuant*, and *Occurrent*. Table 2 shows a segment of the ontology with concept identifier, the description of the class, and mappings to existing ontology classes, which highlights that a broad segment of classes in MDO have been mapped to classes in existing ontologies. However, we noted that classes describing specific PD clinical features are not modeled in existing biomedical ontologies, which demonstrates that MDO is complementary to existing biomedical ontologies although there are a significant number of mappings to existing ontology classes. The mappings to existing ontologies will also facilitate the interoperability of AMP PD with other datasets.

**Table 1:**
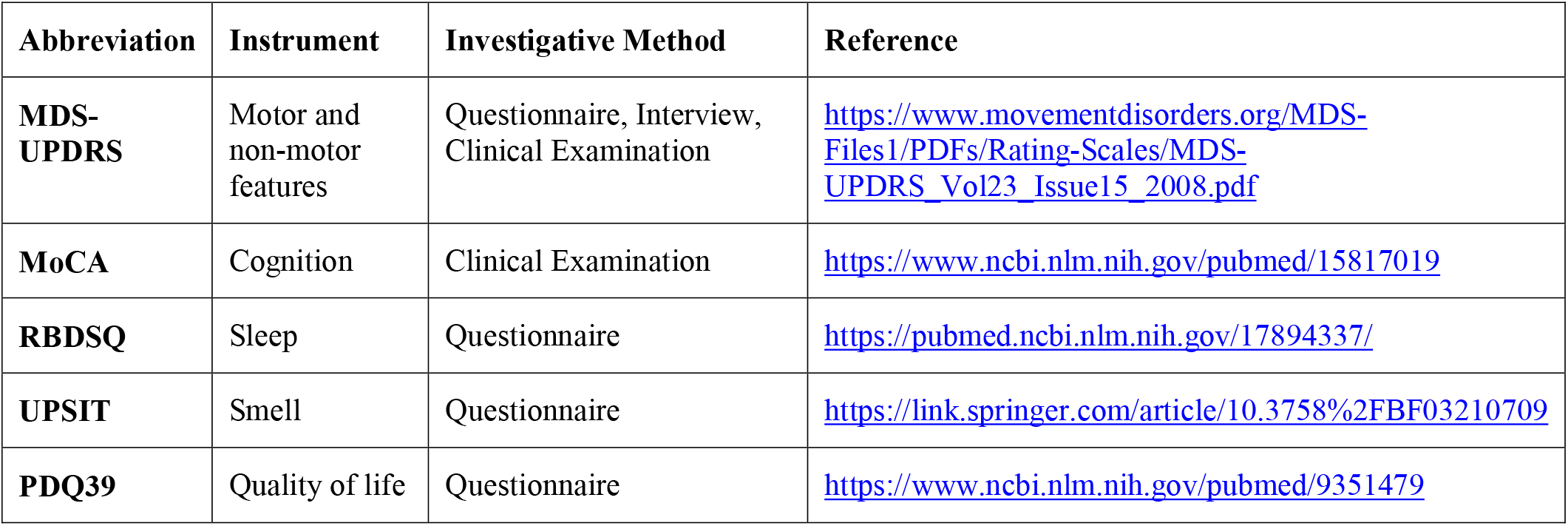
Five types of instruments used in the evaluation of movement disorder patients with method of administering.

**Table 2:** A segment of the Movement Disorder Ontology with mappings to existing ontologies.

| Class ID | Class Description | Mappings to Existing Ontology |
| --- | --- | --- |
| Ataxia (part of Phenomenology class hierarchy) | Loss of muscle coordination | SNOMED CT: Ataxia (CUI: C0004134) |
| Hypoxic (part of the Etiology class hierarchy) | Decrease in level of oxygen in body | NCIT: Hypoxia |
| AlphaSynucleinTest (part of FluidBiomarker class hierarchy) | Laboratory test for alpha synuclein | LOINC: Alpha synuclein<br>Ag:PrThr:Pt:Tiss:Ord:Immune stain |
| Carbidopa (part of the DrugIngredient class hierarchy) | Drug ingredient used in treatment of PD | RxNORM:Carbidopa (CUI: C0006982) |
| DopamineTransporterScan (part of the ImagingModality class hierarchy) | A recently developed imaging modality to aid diagnosis of PD | RCTV2: Dopamine transporter scan (Read Clinical Terminology Version 2) |

Table 3 shows the values for structural metrics used to evaluate MDO. The full ontology consists of 203 classes distributed across the three BFO classes described above. The *DependentContinuant* class has the highest number of classes followed by *IndependentContinuant*. The *Occurrent* class has somewhat lower number of classes; however, we expect a greater number of classes modeled as its subclasses in the next version of MDO as we represent more treatment plans and protocols in MDO. The maximum depth of classes is 7 and the average number of sibling classes is 7 with the highest number of sibling classes modeled as subclasses of *DependentContinuant*. The maximum number of sibling classes are modeled as subclasses of the class *MotorSymptom*. There is a total of 27 classes that are mapped to external ontology classes in the current version of MDO, which is expected to increase as the coverage of terms in MDO increases in the next version of the ontology.

**Table 3:** The structural metrics of the Movement Disorder Ontology.

| Structural Metrics | MDO Total | Dependent Continuant | Independent Continuant | Occurrent |
| --- | --- | --- | --- | --- |
| Classes | 203 | 166 | 29 | 8 |
| Maximum depth of classes | 7 | 7 | 4 | 4 |
| Average number of sibling classes | 7 | 7 | 5 | 2 |
| Maximum number of sibling classes | 12 | 12 | 8 | 3 |
| Classes with single child class | 1 | 1 | 0 | 0 |
| Mappings to external ontologies | 27 | 20 | 6 | 1 |

## Discussion and Conclusions

Pathophysiologically, there has been increasing evidence that PD in its current classification scheme is not really a single disease, but rather a set of different subtypes with varied and yet overlapping etiologies. Alpha-synuclein (α-Synuclein) protein misfolding and accumulation in form of Lewy bodies is considered pathophysiological hallmark of PD. However, PD patients with Leucine-rich repeat kinase 2 (LRRK2) mutations don’t exhibit significant or any Lewy body deposition in neuropathological studies. On the other hand, PD patients with glucocerebrosidase (GBA) mutations have significantly higher Lewy body deposition, compared to patients with no GBA mutations. Beyond these three key pathophysiological players, that is α-Synuclein, LRRK2 and GBA, there are several other genes and proteins which influence and / or lead to PD phenotypes, including Parkinson juvenile disease protein 2 (PARKIN), PTEN-induced kinase 1 (PINK1), Vacuolar protein sorting-associated protein 35 (VPS35), microtubule-associated protein tau (MAPT) / tau, beta-amyloid 42 (Aβ42), etc. In terms of biomarkers, several fluid and imaging biomarkers have been proposed to be associated with PD and APD, however, none have been validated to be sensitive or specific enough for clinical use. Leading candidates include alpha-synuclein, tau, phosphorylated-tau, Aβ42 in cerebrospinal and blood, as well magnetic resonance (diffusion and iron) and nuclear (FDG-PET) imaging modalities. Therefore, there are no established classification scheme for PD that can be used to structure the class hierarchy in MDO to reflect clinical practice.

Currently, MDO only includes clinical and research terms relevant to PD and APD, given our current focus on these disorders. However, several of the existing terms already cover important aspects of other movement disorders. In addition, we are currently expanding MDO to include all relevant clinical and research terms of all movement disorders. In particular, we plan to model the complexity of the underlying pathophysiological characteristics that lead to disparate phenotypes in patient (described above) with an emphasis on modeling genotypic data with explicit correlation with symptoms exhibited by patients. In addition, there is a clear need to model the neuropathology findings in terms of fluid and imaging biomarkers described in the above paragraph.

In conclusion, we presented the development of MDO as an integrated domain-specific ontology for movement disorders with fine level granularity in terms of modeling of clinical features, including phenotypic traits, which are routinely used by movement disorder specialists for diagnosis, progression, and prognosis in patients. The objective of MDO is facilitate ontology-mediated access to high quality data repositories such as AMP PD and support clinical research studies to discover biomarkers or improve treatment plans for modifying the course of PD. We are currently in process of making MDO available at BioPortal (https://bioportal.bioontology.org/) and applying to clinically relevant uses cases.

## Data Availability

Not applicable

## Acknowledgement

This project was supported in part by the Clinical and Translational Science Collaborative (CTSC) of Cleveland which is funded by the National Institutes of Health (NIH), National Center for Advancing Translational Science (NCATS), Clinical and Translational Science Award (CTSA) grant, UL1TR002548. The content is solely the responsibility of the authors and do not necessarily represent the official views of the NIH. This project was also supported in part by the NSF grant# 1636850.

